# Electronic health records reveal variations in the use of blood units by hour and medical specialty

**DOI:** 10.1101/2024.11.08.24316743

**Authors:** Esa Turkulainen, Elissa Peltola, Markus Perola, Miika Koskinen, Mikko Arvas, Minna Ilmakunnas

**Affiliations:** Finnish Red Cross Blood Service, Research and development; Helsinki University Hospital, IT Management; Finnish Institute for Health and Welfare (THL); Helsinki University Hospital, Diagnostic Center; Department of Anesthesiology and Intensive Care Medicine, Helsinki University Hospital and University of Helsinki

## Abstract

**BACKGROUND AND OBJECTIVES:** Efficient blood supply chain requires accurate demand estimates. Blood demand is created by clinicians making transfusion decisions based on patient status. To better understand the use of blood units, we tracked their use hourly and across a large hospital organization.

**MATERIALS AND METHODS:** We analyzed blood use in adult patients over 2021-2022 at HUS Helsinki University Hospital, serving a population of 1.7 million and consuming a third of blood units used in Finland. We utilized electronic health records (EHR) to map transfusions to patient demographics, diagnoses, medical specialties, treatment events, surgical procedures, and laboratory values. Data were matched to transfusion of red blood cells, platelets and plasma using timestamps and treatment episodes.

**RESULTS:** In total, 107,331 units were transfused to 19,637 unique patients in 50,978 transfusion episodes. Most transfusions occurred in emergency settings, with 61.5% of use driven by emergency department admissions. The most common diagnoses were malignant neoplasms, anemias, and cardiovascular diseases. In total, 47.9% of transfusions were associated with a surgical procedure. Of these, 72.9% were for urgent surgery. Blood use peaked in the early evening and was lowest during morning office hours.

**CONCLUSION:** The study offers a comprehensive picture of blood use in one of the largest European hospital organizations. In addition to elective use, a significant portion of blood demand is driven by urgent and emergency needs, which introduces some uncertainty in predicting blood use. Future studies should aim to understand both elective and emergency blood use to help improve demand estimates.

**Highlights:**

- This study presents the first investigation into intraday blood use within a large hospital district, with precise transfusion timestamps enabling alignment with patient events.
- Over the past 25 years, blood demand has shifted from surgery to hematology in the HUS Helsinki University Hospital.
- The majority of blood is used in emergencies and urgent care, mostly outside office hours.

## INTRODUCTION

An efficient blood supply chain satisfies demand without wasting any blood products. Most research on blood supply chains has focused on storage and logistics, assuming demand either as a constant or as a distribution.^1^ Research on the demand has relied on autoregressive time series analysis, sometimes assisted by machine learning.^2–4^

Transfusion decisions by clinicians define blood demand. Blood supply chains, predominantly managed by independent blood services, are frequently decoupled from the actual demand within hospitals. Tackling this barrier by studying when and how blood is used in hospitals can improve the efficiency of blood supply chains in the future. Several studies have addressed blood use in hospitals, aims varying from surveying transfusion practices,^5–29^ to estimating long-term future demand,^5,6,12,24,30–40^ and understanding indications for short-term demand.^41^ In these studies, patient demographics, diagnoses, medical specialties, hospital departments, operation types, number and type of transfused products, blood loss and hemoglobin measurements, and mortality have been used variably to characterize blood utilization. However, a comprehensive analysis integrating these variables within a large-scale, real-world hospital setting is needed to provide actionable insights for optimizing blood supply management.

We analyzed blood use in Helsinki University Hospital (HUS). HUS uses a third of the over 200,000 blood products distributed annually by Finnish Red Cross Blood Service (FRCBS), the sole operator collecting and distributing blood products in Finland. We aimed to build a comprehensive picture of blood demand in HUS by utilizing electronic health records (EHR) to delineate when, where, and how blood is used. Understanding the patterns of blood use in the largest hospital in the country could help approximate nationwide blood demand and support future blood collection planning.

## STUDY DESIGN AND METHODS

The study population comprised adult (≥18 years) patients in HUS who received transfusions during 1.1.2021-31.12.2022. HUS has a population base of approximately 1.7 million, representing 30% of the Finnish population. The time period reflects the usual blood use in HUS after the initial phase of the COVID-19 pandemic and the implementation of a new EHR system at 2019-2020. The data was extracted by HUS ICT Management from HUS data lake and delivered pseudonymized into a secure analytics platform, HUS Acamedic. All data were drawn from EHR and clinical in nature, with no access to a blood bank laboratory information management system (LIMS). The data were subsequently filtered and crossmatched by patients and transfusions to match individual patients and their records. A schematic of this process is presented in **Figure 1**.

**Figure 1:**
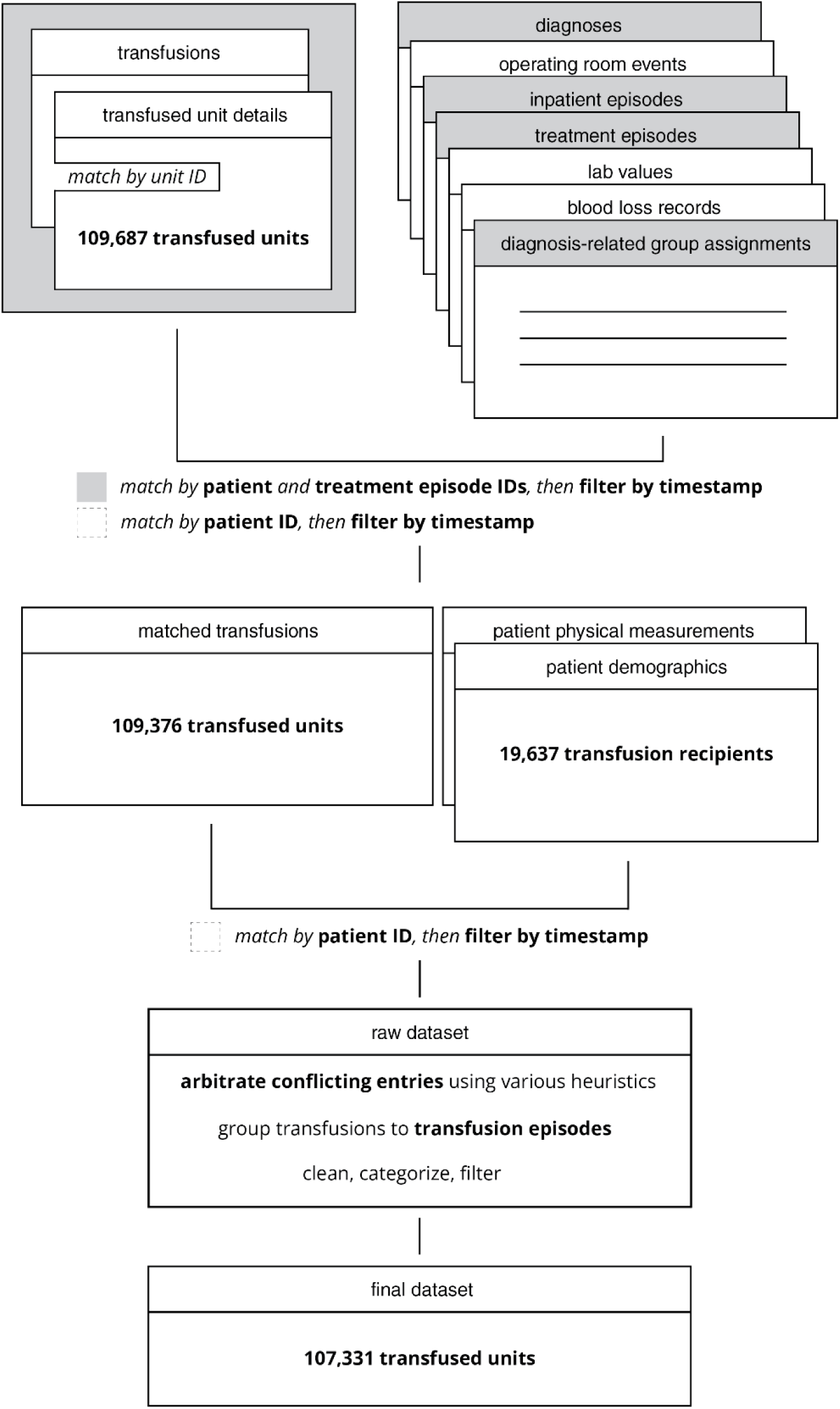
Summary diagram of dataset compilation process.

The institutional review board at HUS granted a research and data permit for this study (HUS/579/2022, 17.10.2022). As the study is based purely on registry data, obtaining ethics committee approval was unnecessary pursuant to the Act on the Secondary Use of Health and Social Data (26.4.2019/552) in Finland.

### Transfused units and transfusion episodes

Transfused units were identified using a unit identifier and a timestamp indicating either the beginning or the end of its transfusion. When either of these timestamps was missing, we approximated the event times computationally (**Supplement Section S1.1)**.

To cluster units related to the same procedure or treatment episode, we defined transfusion episodes as sequential transfusions for the same individual with no more than 3 hours between the end of a transfusion and the start of the next transfusion. The 3-hour limit was chosen after exploratory data analysis of transfusion time differences (**Supplement Section S2.7**).

### Primary diagnosis and medical specialty

To estimate the diagnosis necessitating transfusion, we applied hierarchical matching criteria. Primary diagnoses were retrieved from events closest to the start of the transfusion unless postoperative diagnosis was available. All diagnoses were coded using the International Classification of Diseases, Tenth Revision (ICD-10), Finnish edition. To further characterize reasons for transfusion, we used Diagnosis Related Groups (DRG) from the NordDRG classification.^42^

We mapped transfusions to the medical specialty treating the patient at transfusion. The specialties follow the coding of the national Care Register for Health Care in Finland. We further classified specialties into operative and conservative (**Supplement Section S1.2**).

### Patient presentation and primary location

We classified the various patient admittance channels into emergency/urgent, scheduled treatment, transfer between departments or hospitals, and other unspecified routes.

Patient location within HUS (unit or department) was classified into hospital ward, OR, emergency department (ED) including prehospital emergency medical system, intensive care unit, outpatient unit, dialysis unit, endoscopy unit, and obstetric unit. Some units were classified as “Other” due to their low prevalence in the data. The location for transfusion was determined with the timestamp proximity method.

### Operating room events

Transfusions starting within six hours before surgery were categorized as preoperative, during surgery as intraoperative, and within one week afterwards as postoperative if the patient remained hospitalized.

Operation urgencies follow established national classifications^43^, and were assigned by the surgeon booking the procedure (for emergencies in <2 h, in <6 h, in <24 h, in <48 h, and within one week; for elective in 30, 90, and 180 days). Organ transplant and special requirement surgeries were separate classes.

Procedures follow the NOMESCO Classification of Surgical Procedures (NCSP).

### Hemoglobin levels and blood loss

We matched latest (up to 24 hours) pre-transfusion and earliest (up to 2 days) post-transfusion hemoglobin (Hb; g/L) measurements to red blood cell (RBC) transfusions, when available. If multiple measurements shared a timestamp, the average of these measurements was taken.

Pre-episode bleeding was calculated as the cumulative blood loss within 24 hours before the transfusion episode or until the end of the previous episode to prevent overlap. To calculate relative blood loss, we estimated patient blood volume using Nadler’s equation (**Supplement Section S1.4**).

Similarly, we matched platelet counts (×10^9^/L) to platelet transfusions and International Normalized Ratios (INR) to plasma transfusions (OctaplasLG). For the analysis of plasma transfusion threshold, we excluded patients undergoing plasma exchange. Missing INR measurements were estimated using prothrombin time (PT) if available (**Supplement Section S1.3**).

### Statistical analysis

Data are presented as number (%) or median (interquartile range, IQR). Distributions for imputation and visualization were estimated using kernel density estimation.

All development code has been made available on Github.^44^

## RESULTS

The unprocessed transfusion data contains 74,832 RBC units, 15,121 platelet units, and 17,274 plasma units transfused to adult patients between 2021 and 2022. This covers 76.7%, 72.1%, and 71.1% of RBC, platelet, and plasma units delivered to HUS between 2021 and 2022, respectively, according to FRCBS. Reasons for incomplete coverage include e.g. exclusion of children from analysis, the expiry of units (sold to HUS but not transfused; data on expired units not available), and incorrect or incomplete data entry.

We further excluded 1,902 units from the analysis, because their descriptive statistics made some patients plausibly identifiable.

Our final data set comprises 19,637 unique adult patients with 50,978 transfusion episodes. Most recipients were female (56.7%), while most units were administered to males (53.8%). The patients received 107,331 units in total: 74,805 RBC units (of which 3,772/5.0% radiated and 1,378/1.8% issued for emergency transfusion), 15,090 platelet units, and 17,276 OctaplasLG units. The general characteristics of transfusion recipients are presented in **Table 1**. Most of the transfusion episodes involved only one type of blood component, usually RBCs (**Figure 2**). The most frequent multicomponent transfusion comprised RBC and platelets. The median number of RBC and platelet units transfused was two per transfusion episode, with 49.5% and 17.7% transfused as a single unit, respectively. The proportion of single unit transfusions was higher (RBC 52.8% and platelets 27.7%) when transfusions of single units of either RBC or platelets concomitantly with other blood products (i.e. as a part of multicomponent transfusion) were included (**Supplement Section S2.3**). For plasma, slightly over 50% of units were transfused as batches of eight or more units, mainly for plasma exchange. Most RBC transfusions were ABO-identical (**Supplement Section S2.6**).

**Figure 2:**
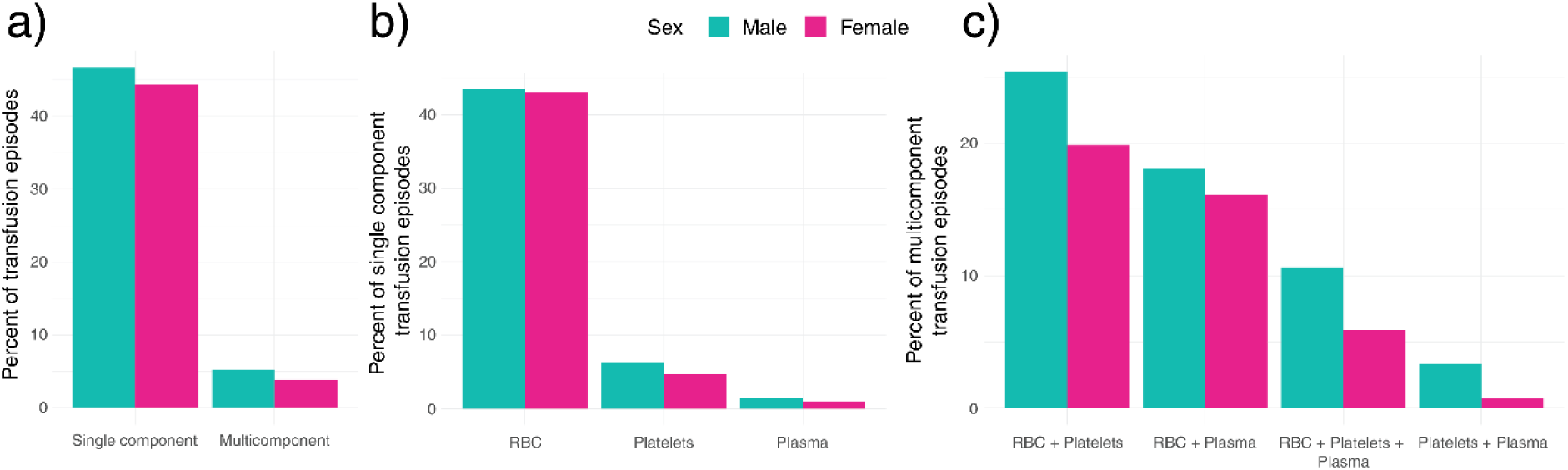
a) Proportions of single vs. multicomponent transfusion episodes. b) Proportions of components in single component transfusion episodes. c) Proportions of component combinations in multicomponent transfusion episodes. Figure represents 99.999% of the dataset.

**Table 1:** Descriptive statistics of transfused blood products. Medical specialties follow the Finnish national classification.

|  | Overall | Female | Male |
| --- | --- | --- | --- |
| **Number of transfusions (% of total)** | 107,331 (100) | 49,535 (46.2) | 57,795 (53.8) |
| **Number of transfusion episodes (% of total)** | 50,978 (100) | 24,558 (48.1) | 26,419 (51.8) |
| **Number of unique patients (% of total)** | 19,637 (100) | 11,130 (56.7) | 8,506 (43.3) |
| **Age at transfusion, N (% of group)** |  |  |  |
| 18–29 y | 1,168 (5.9) | 894 (8) | 274 (3.2) |
| 30–49 y | 3,270 (16.6) | 2,402 (21.5) | 868 (10.2) |
| 50–69 y | 5,480 (27.8) | 2,456 (22) | 3,024 (35.4) |
| 70–89 y | 8,608 (43.7) | 4,567 (40.9) | 4,040 (47.3) |
| >90 y | 1,195 (6.1) | 850 (7.6) | 345 (4) |
| **Median age (IQR)** | 69 (53–79) | 70 (42–80) | 70 (58–78) |
| **Transfusions per patient, median (IQR)** |  |  |  |
| Red-cell units | 2 (2, 4) | 2 (2, 4) | 3 (2, 5) |
| Platelet units | 2 (2, 4) | 2 (2, 4) | 2 (2, 4) |
| Plasma units | 3 (2, 5) | 3 (2, 4) | 3 (2, 6) |
| **Blood loss (ml) per transfused unit, median (IQR)\*** | 300 (125, 550) | 302 (150, 625) | 239 (100, 450) |
| **Length of inpatient stay in days, median (IQR)** | 1.3 (0.4, 5.1) | 1.38 (0.4, 4.9) | 1.24 (0.4, 5.7) |
| **Medical specialty of recipient, N units (% of group)** |  |  |  |
| Emergency medicine | 14,561 (13.6) | 7,352 (14.8) | 7,209 (12.5) |
| Hematology | 11,285 (10.5) | 5,105 (10.3) | 6,180 (10.7) |
| Orthopedics and traumatology | 8,609 (8.0) | 4,901 (9.9) | 3,708 (6.4) |
| Gastrointestinal surgery | 8,241 (7.7) | 3,238 (6.5) | 5,002 (8.7) |
| Gastroenterology | 8,215 (7.7) | 3,337 (6.7) | 4,878 (8.4) |
| Cardiac surgery | 8,108 (7.6) | 2,055 (4.2) | 6,053 (10.5) |
| Nephrology | 6,965 (6.5) | 3,083 (6.2) | 3,882 (6.7) |
| Vascular surgery | 6,439 (6.0) | 1,988 (4.0) | 4,451 (7.7) |
| Obstetrics | 4,532 (4.2) | 4,532 (9.2) | ≤ 5 (0) |
| Internal medicine | 3,972 (3.7) | 1,622 (3.3) | 2,350 (4.1) |
| General internal medicine | 3,797 (3.5) | 1,758 (3.6) | 2,039 (3.5) |
| Plastic surgery | 2,952 (2.8) | 1,168 (2.4) | 1,784 (3.1) |
| Transplant surgery | 2,913 (2.7) | 1,059 (2.1) | 1,854 (3.2) |
| Gynecology | 2,563 (2.4) | 2,563 (5.2) | ≤ 5 (0) |
| Urology | 2,295 (2.1) | 595 (1.2) | 1,700 (2.9) |
| Other | 3,371 (3.1) | 1,458 (2.9) | 1,913 (3.3) |

### When and where is blood needed?

The highest demand occurred in the late afternoon, peaking at around 6 PM. The lowest demand was observed in the morning (**Figure 3A**). We also examined blood use within a calendar year as weekly series. Blood use for elective, but not for urgent, surgery decreased during the summer. In conservative specialties, blood use remained somewhat constant throughout the year (**Supplement Section S2.1**).

**Figure 3:**
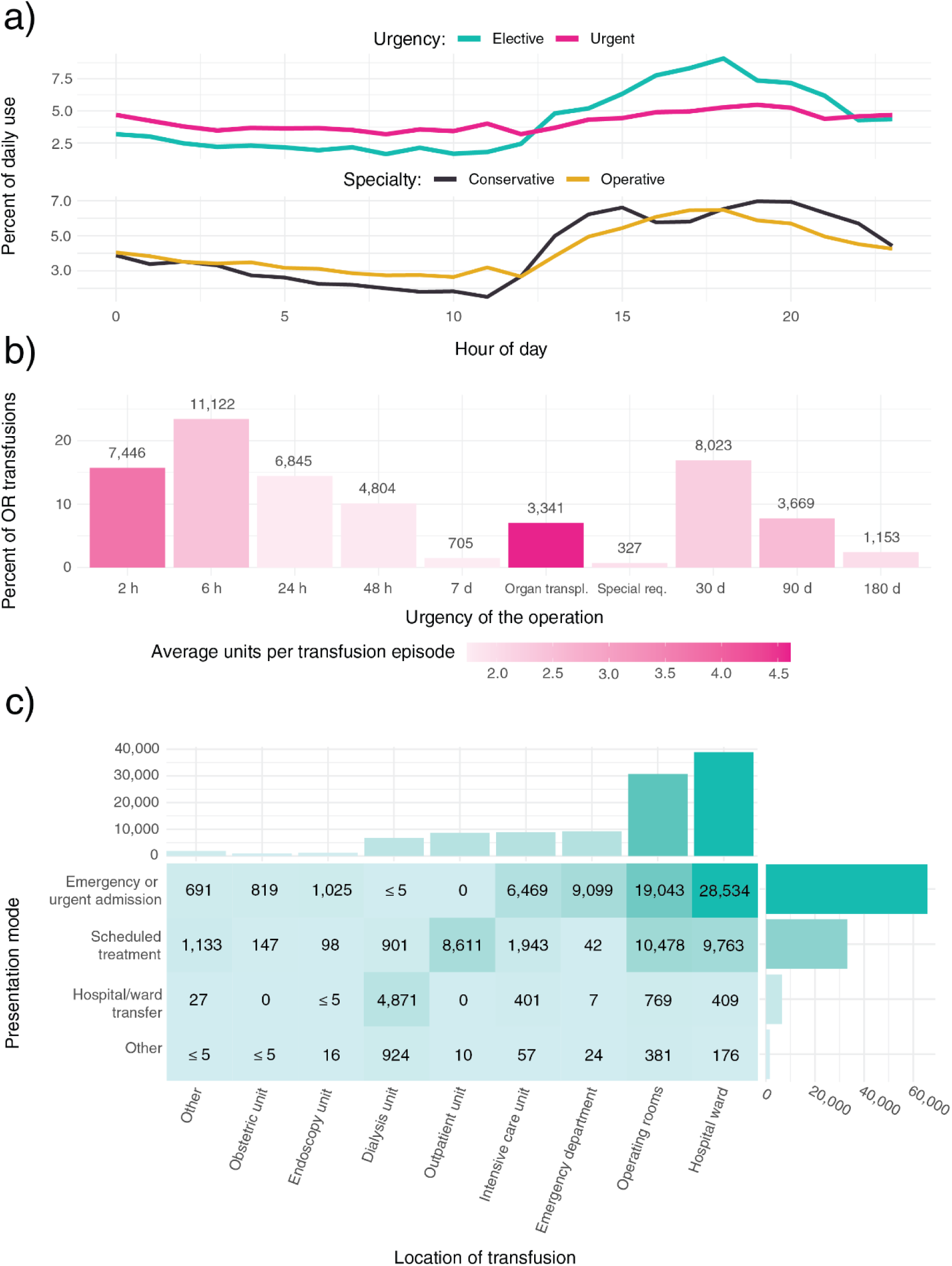
a) Intraday blood use in operating rooms (OR) by urgency (elective vs. urgent) and in total by medical specialty (conservative vs. operative). Urgency categories were available for 44.2% of the dataset and medical specialties 99.97% of the dataset. The y-axis represents the proportion of total daily use. b) OR transfusions by urgency. Category labels indicate the time period in which the patient should be operated. Categories between “2 h” and “7 d” are urgent; categories “30 d”, “90 d”, and “180 d” are elective. Special categories: “Organ transpl.” = organ transplantation and “Special req.” = operations requiring special resources. Color gradient indicates how many units on average are transfused together. c) Heatmap of transfused units between patient presentation method and transfusion location within the hospital. Figure represents 99.58% of the dataset.

**Figure 3B** shows the distribution of transfusions associated with OR events, with 53.6% of units used in urgent operations within 24 hours, and only 27.1% for elective surgery. Transfusions related to organ transplantation comprised 7% of OR transfusions and 3.1% of all transfusions. Finally, 15.5% of transfused units were used in massive transfusions (episodes with at least 10 transfused units).

The heatmap in **Figure 3C** details transfusions based on the patient’s mode of presentation and the location within the hospital at transfusion. Most transfusions were administered for patients admitted through the ED, and while the ED was a common location for transfusions, most patients received their transfusions mostly in the wards and OR.

### Why is blood needed?

The single largest ICD-10 diagnosis categories for blood use were malignant neoplasms (codes C00-97), aplastic and other anemias (D60-64), and diseases of arteries, arterioles, and capillaries (I70-79) (**Figure 4**). The median time from a diagnosis to the first transfusion associated with the same diagnosis was highly variable (**Supplement Section S2.2**).

**Figure 4:**
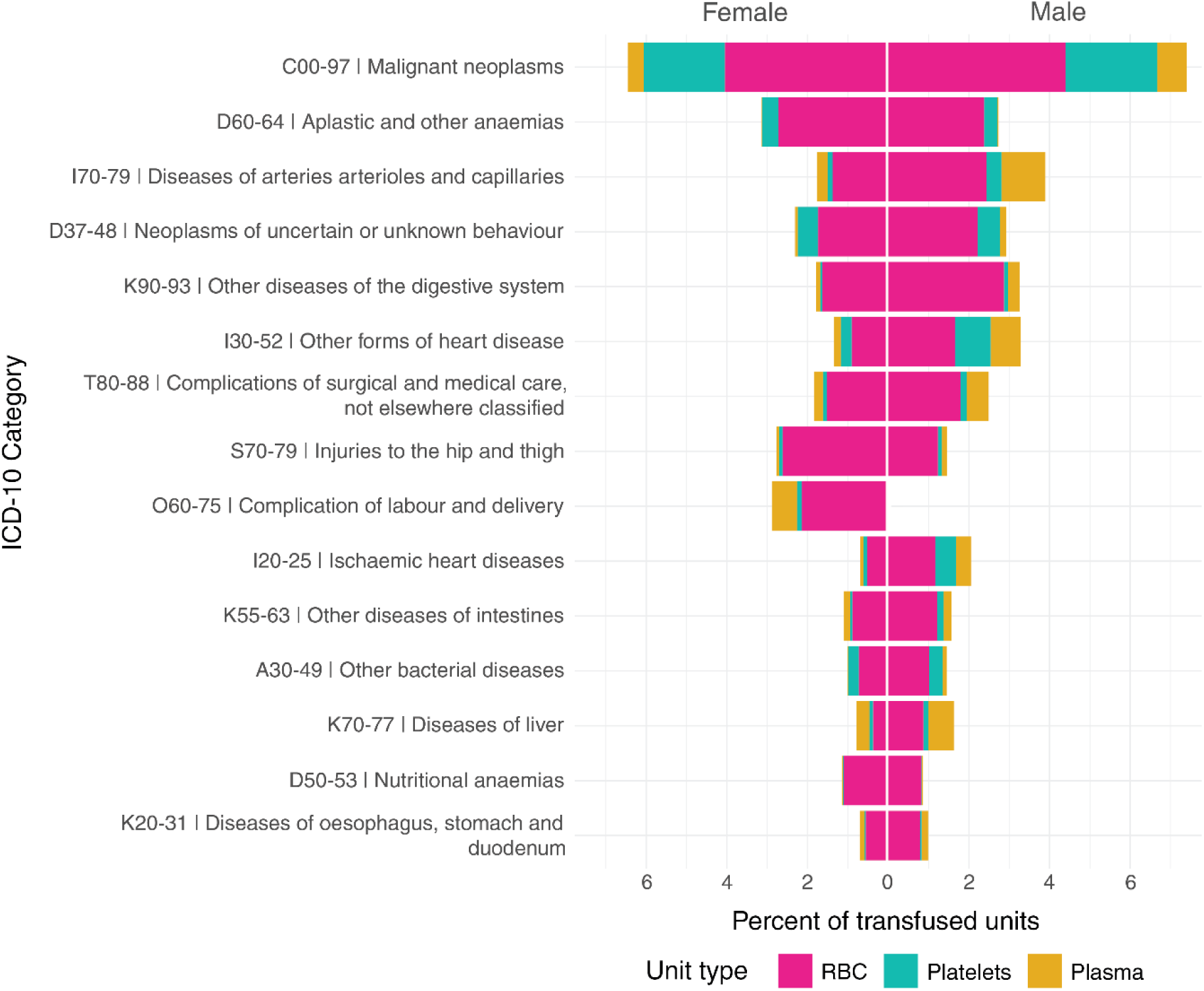
Fifteen most frequent ICD-10 categories associated with transfusions. Ranking was computed from 99.67% of the dataset, and the figure represents 65.65% of the dataset.

Grouping transfusions by DRG classification showed various hematologic diseases, gastrointestinal hemorrhage, liver transplantation, and delivery with complications requiring transfusions most often (**Figure 5**).

**Figure 5:**
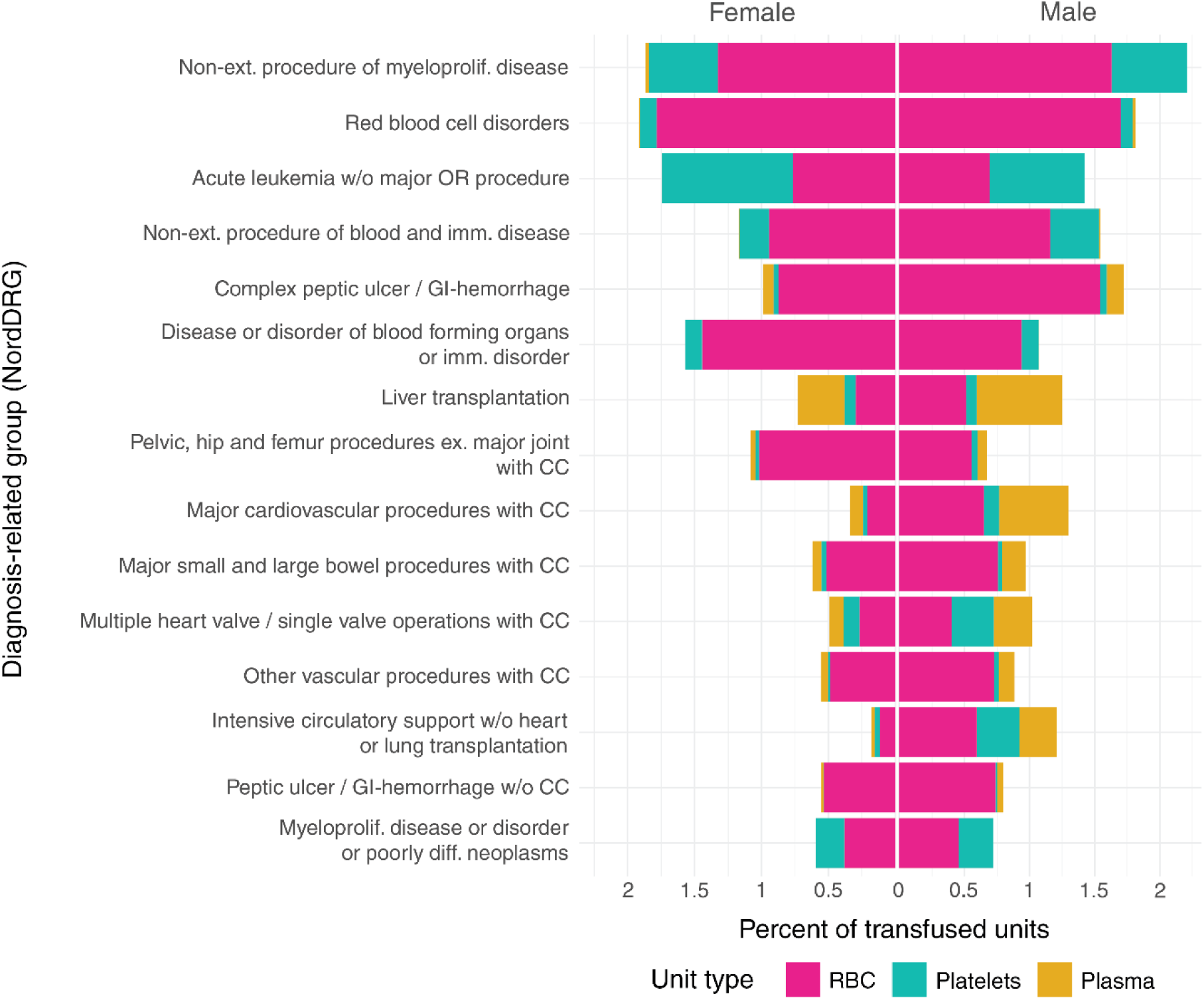
Fifteen most frequent diagnosis-related groups (NordDRG) associated with transfusions. Non-ext. = non-extensive, myeloprolif. = myeloproliferative, w/o = without, imm. = immunological, GI = gastrointestinal, ex. = excluding, CC = complications. Ranking was computed from 91.40% of the dataset and the figure represents 32.99% of the dataset.

Most transfusions were not associated with operations (52.0%). Most (83.3%) of these were administered in conservative specialties (**Figure 6**). In operative specialties, 15.4% of the transfusions were not directly associated with any procedure. Of the transfusions mapped to operations, 5.0% were administered preoperatively, 21.4% during the operation and 73.6% postoperatively within one week after the operation.

**Figure 6:**
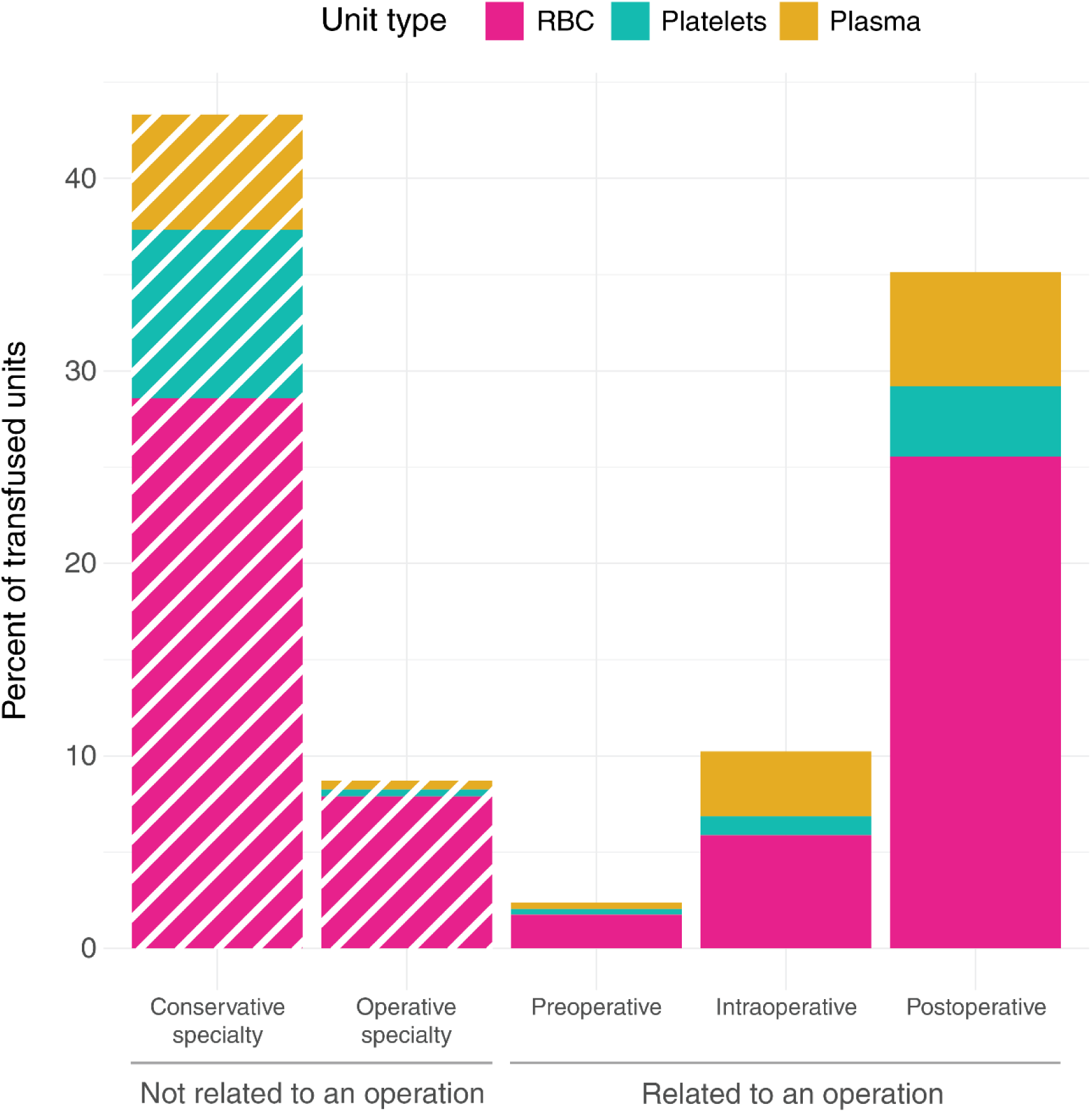
Transfused blood products associated with specific operations. Products not associated with an operation were classified into conservative and operative specialties. Preoperative = transfusion starting within 6 hours before the operation (preoperative), intraoperative = during the operation, postoperative = within a week after an operation. The y-axis represents the percentage of all transfused units. Figure represents 99.81% of the dataset.

Procedures of the hip joint and thigh (NF codes in NCSP) were associated with the most blood use. Together with procedures after delivery or abortion (MB), and procedures of the liver (JJ), they account for 10.2% of all blood used in HUS (**Figure 7**).

**Figure 7:**
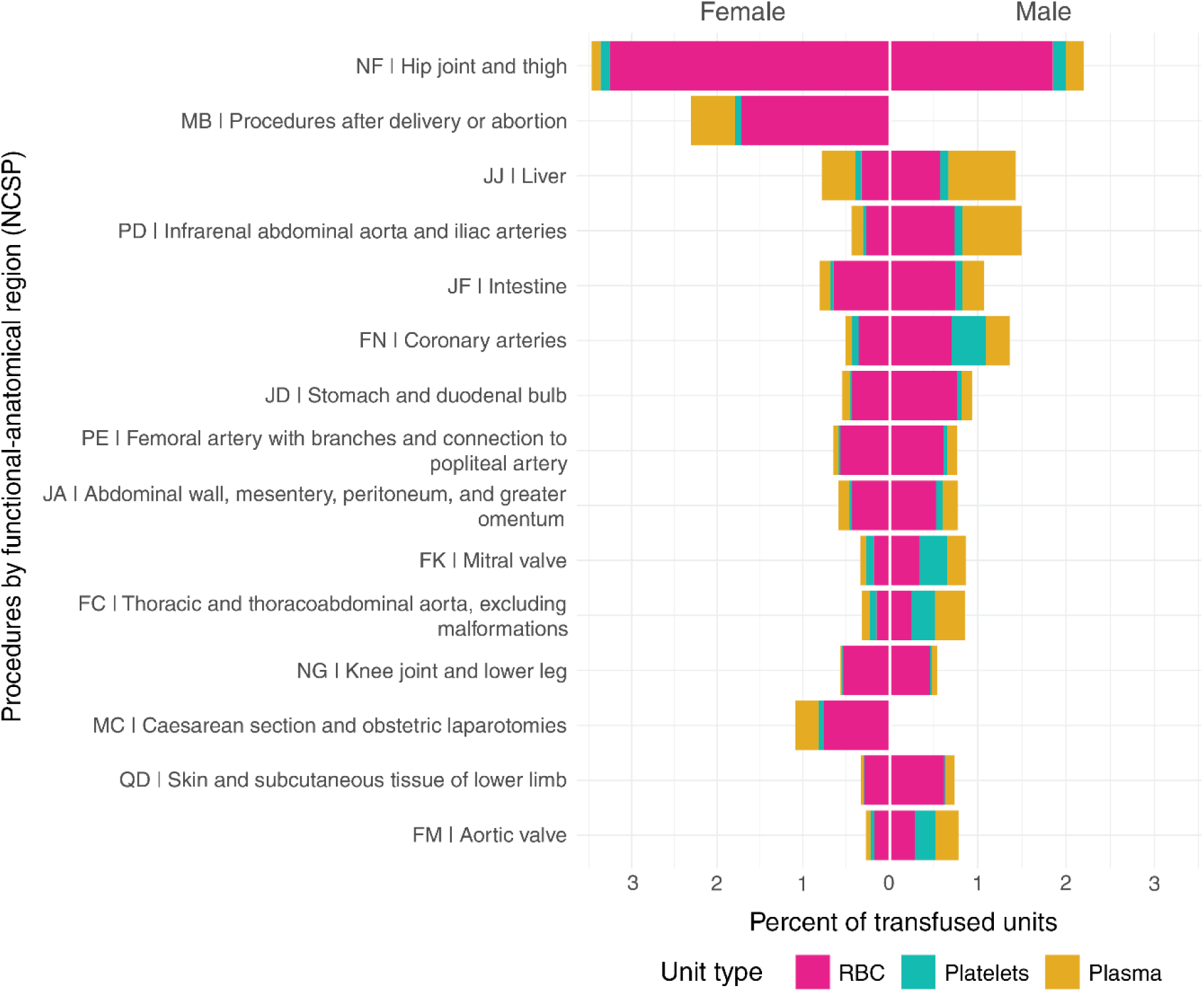
Fifteen most frequent operation types (by NCSP, NOMESCO Classification of Surgical Procedures) associated with transfusions. The x-axis represents the percentage of all transfused units. Ranking was computed from 43.79% of the dataset, and the figure represents 26.78% of the data.

The median pre-transfusion Hb was 80 g/L, with very little variation between sexes (79 (79-80) g/L;80 (80-80) g/L, and 80 (80-80) g/L for premenopausal women, postmenopausal women, and men, respectively). While the mode of pre-transfusion Hb was below 80 g/L for all sex groups, the distributions for men and postmenopausal women exhibited a secondary peak spanning 80-89 g/L, suggesting a distinct subset of patients receiving RBC transfusions at higher Hb levels. We compared transfusions at the peak range (70-79 g/L) to transfusions at the secondary peak range (80-89 g/L) by patient age group, main diagnosis, and main procedure, and found that patients at the higher pre-transfusion Hb range tended to be older and more commonly underwent surgery (56% vs. 42%), typically for peripheral arterial disease or femoral fractures (**Supplement Section S2.8**).

The median increase in Hb per administered unit was largest for postmenopausal women (+7 g/L, +9.6 g/L, and +7 g/L, respectively). When a transfusion could be mapped to blood loss, women generally bled more relative to their estimated blood volume per transfused unit (medians 11.6%, 6.0%, and 4.4% for premenopausal women, postmenopausal women, and men, respectively) (**Figure 8**). The median blood loss was 302 (150-625) ml in women and 239 (100-450) ml in men. Men on average required more platelets and OctaplasLG (**Figures 4, 5, and 6**).

**Figure 8:**
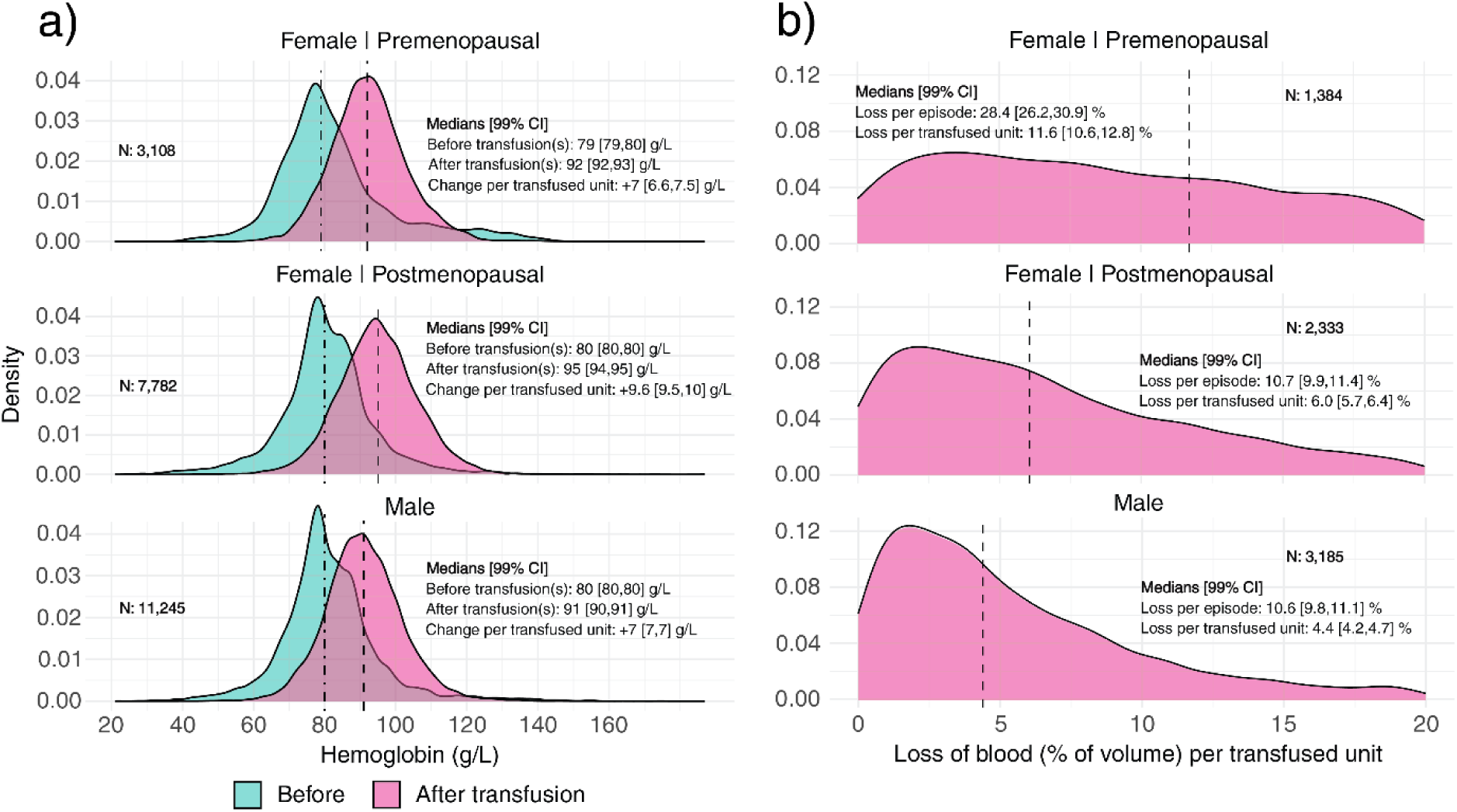
a) Distributions of hemoglobin measurements. Only patients with both pre- and post-transfusion measurements were included. Figure represents 35.68% of the data. b) Distributions of blood loss before and during a transfusion episode, as proportion of recipient total blood volume per unit of transfused blood. Figure includes only recipients for whom any blood loss was reported and represents 22.27% of the data.

For platelets, the median transfusion thresholds were 66 (35-94) ×10^9^/L in women and 76 (42-121) ×10^9^/L in men. For plasma, the thresholds were INR 1.3 (1.1-1.6) in women and 1.4 (1.3-1.8) in men, respectively (**Supplement Section S2.5**).

## DISCUSSION

Although several studies have addressed transfusion practices during the past 20 years, most have captured only a relatively small number of transfusions^5,12,14,16,17,38^ or transfused patients^13,36,41^, while some have included only a short time period^10,14,17,26,27,32,36,37,40^ or a specific patient population^16,19,26,28,31,41^. Several studies looked only at the use of RBCs^2,11,14,17,25,34,37,40^ or platelets^7,10,23^. Further, older studies may not reflect contemporary transfusion practices and thus the current blood use. To overcome these limitations, we examined 107 331 units of any blood units transfused to 19 637 recipients in HUS between 2021 and 2022.

To our knowledge, the current study is one of the first to examine the intraday blood demand patterns. By looking at hourly demand, we noticed that the daily peak both in blood demand and use occurred outside the office hours of full staffing both in the hospital blood bank and hospital units transfusing the blood. Further, the intraday demand was similar in conservative and operative specialties. This evening peak in blood demand has severe implications to patient safety. A disproportionately high number of laboratory errors related to pre-transfusion testing occur outside the office hours.^45,46,47^ Patient identification errors and incomplete transfusion documentation are more common, and adequate clinical monitoring during transfusion is often neglected.^48^ These errors can lead to transfusing wrong components^45,46^ and poor recognition of transfusion reactions.^48^ Our findings underline the importance of adequate staffing during on-call hours. Any non-urgent transfusions should be avoided during on-call hours and delayed to daytime. Further, nursing personnel should be encouraged to adhere strictly to the safe transfusion practices even at nighttime.

A plausible explanation for the observed evening peak in blood demand is transfusion for acute bleeding and emergency surgery. Previous studies have examined blood use by admission type or specifically in elective surgery or EDs^11,17,26,38,41^. However, less is known about blood use between emergency/urgent admissions versus scheduled treatment, and the urgency of transfusions in these patients^21,31,40^. Internationally, 15.9-82.3% of blood requests are for emergencies or non-elective surgery.^25,36,49^ In our study, 61.5% of blood use was driven by patients presenting to the ED. Further, when administered in the OR, only 27.1% of transfusions were for elective operations. This high frequency of transfusions for emergency/urgent patients has both clinical and blood banking implications. Measures aimed at reducing transfusions, especially adequate treatment of anemia, are more effective when started early before operations.^50^ This may be impossible in emergency/urgent patients, but specific treatments aimed at the underlying cause of anemia should be started promptly to avoid overuse of blood products. For blood supply chain management, understanding the variance in demand is a prerequisite for developing robust predictive models.

Although the majority of blood demand was for urgent admissions and predominantly elderly recipients, an analysis of ICD-10, DRG, and NCSP categories revealed a complex profile of blood use. ICD-10 codes related to neoplasms, anemias, and cardiovascular diseases accounted for 40% of blood usage, largely consistent with previous findings.^6^ Examination of DRG codes confirmed that several of the largest patient cohorts requiring transfusion were hematological. This represents a significant shift, as 25 years ago several surgical specialties used more blood than hematology^49^, and likely reflects decreased intraoperative bleeding with improved perioperative care and surgical techniques. Indeed, the majority of transfusions directly associated with specific procedures were postoperative, i.e. within a week after the operation, even when most transfusions were for patients initially admitted as emergency/urgent cases.

A previous audit of transfusion practices in Finland reported a median pre-transfusion Hb of 82 g/L.^20^ In our mixed cohort of both bleeding and non-bleeding patients, the median pre-transfusion Hb was 79-80 g/L, suggesting a declining trend. In half of the patients, the threshold was higher than the recommended 70-80 g/L.^51,52^ These patients were typically elderly surgical patients. Similar to hemoglobin, pre-transfusion platelet counts were higher and pre-transfusion INR levels were lower than those recommended in various guidelines.^53,54^ Our results may be explained by the significant proportion of transfusions related to emergency/urgent admissions or corresponding surgery, when strict adherence to restrictive transfusion thresholds may not be clinically feasible or the actual recommended transfusion thresholds, especially for platelets, are higher.^50^ Similarly, the emergency/urgent case profile may partly explain the relatively low number of single unit transfusions. While the median number of RBC units received was two, the most frequent transfusion episode (all sequential transfusions that start within 3 hours after the end of the previous transfusion) consisted of a single RBC unit (49.5%), lower than the previously reported 62-64% in similar hospital systems.^29,49^ Our results underline the importance of continuing follow-up of transfusion practices and feedback to the clinical staff to ensure the appropriate use of blood products.

While our findings contribute to a more comprehensive understanding of blood utilization, this study has several limitations. The validity of our results is partly contingent on the reliability of data collection and filtering processes and event matching decisions. Although the EHR in HUS provides high-resolution information on patient events, they are prone to systematic errors. Matching transfused units with patient events using time stamps introduces potential inaccuracies into the dataset, a challenge documented previously.^55^ We have therefore strived for transparency in data filtration and matching processes and published our development code on Github.^44^ Our transfusion data coverage in relation to the number of blood products delivered to HUS is incomplete, mostly explained by the exclusion of pediatric population from analysis. We had no access to the discard rate at the hospital blood bank due to unit expiry, an important metric in the assessment of blood supply chain management. Moreover, the generalizability of our findings may be limited by the relatively brief study period and the potential residual effects of the COVID-19 pandemic on blood use. Lastly, a universal definition for transfusion episode is lacking. We allowed maximum 3 hours between transfusions during an individual episode. This time limit, based partly on the local turnaround time for laboratory tests, would allow for assessing the response to transfusion in a non-bleeding patient before making a new transfusion decision independent from the previous transfusion. Likewise, our definitions for pre- and postoperative transfusions are arbitrary and based on clinical feasibility.

The strengths of the study lie in the comprehensive data coverage across different specialties and hospital units. Further, unlike most previous studies, we had access to the detailed data on the mode of patient presentation, the urgency of surgical procedures, and most importantly, the electronically recorded exact timing of transfusions.

Our findings demonstrate that while the majority of blood demand is driven by ED admissions and urgent surgical procedures, thereby complicating the predictability of blood use, a substantial portion of this demand can be attributed to identifiable diagnostic, procedural, and demographic cohorts. This suggests that, despite the inherent variability, hospital EHR possess significant potential for both identifying areas for clinical improvement and enhancing blood demand estimation and optimizing supply chain management.

## ACKNOWLEDGMENTS

We thank the clinicians in HUS for providing valuable comments on the design of this study. MD, PhD Katja Salmela is acknowledged for elucidating hospital blood bank processes, and MD, PhD Riina Jernman for helping to interpret the data on obstetric patients.

This study was supported by Government Research Funding (VTR) and the Finnish Red Cross Blood Service Research Fund as part of the Modeling and predicting blood product demand in HUS Helsinki University Hospital (HUS-RBC) research project.

All authors contributed towards designing this manuscript. The manuscript was written by ET, EP, and MI. Data request was submitted by MI. Data was processed and analyzed by ET and EP.

## Data availability

No data is available outside the authorized use within the HUS Acamedic platform.

