## Supplementary Materials for "Electronic health records reveal variations in the use of blood units by hour and medical specialty"

### S1 Details of data processing

#### S1.1 Imputation of missing transfusion start and end times

The transfusion data contained records of units with and without time stamps for transfusion. In our data processing we have assumed that units with timestamps have been transfused and units that are missing timestamps have been ordered or prepared but not actually transfused. If only one of the timestamps was present, we had to impute the missing timestamp. When the transfusion end time was missing (N = 7), we assumed non-systematic data entry error, and sampled the distribution of transfusion durations across all transfusions and computed the missing end time as start time + sampled duration. When the transfusion start time was missing (N = 671), we assumed (by expert assessment) that most of these were due to emergency transfusions, for which logging the start of the transfusion was not possible. Hence, when sampling transfusion durations, we sampled from the distribution of extremely urgent operations instead of all transfusions. In both cases, distributions were estimated using kernel density estimation.

#### S1.2 Classification of operative and conservative specialties in Finland

Specialties follow the classification used in Finnish

Find code on GitHub!

Care Register (HILMO)[1] for all secondary and tertiary inpatient (hospitalizations and procedures) and outpatient care, including scheduled and emergency care specialist visits. Specialties were further classified dichotomously into conservative and operative by the authors (Table T1.2).

#### S1.3 Estimating missing INR values using PT measurements

Pre- and post-transfusion International Normalized Ratio (INR) measurements were mapped to transfusion episodes using 5-day and 2-day cutoffs, respectively, mirroring the approach used for hemoglobin (Hb) measurements. If Prothrombin time (PT) measurement was temporally closer to the start or end of a transfusion episode than an INR measurement, we calculated the INR using the PT value. The logarithm of INR was approximated using second-order nonlinear regression with logarithmic transformations on input data. For fitting the model, we utilized PT measurements that had corresponding INR values at the same time point. Given that PT is a continuous variable while INR is discretely paced, we calculated mean PT values for each INR value and used the mean PT as an explanatory variable. The model's performance, assessed using 5-fold cross-validation

Table T1.2 | Conservative and operative medical specialties

| | | |
|---|---|---|
| 10 | Internal medicine | Conservative |
| 10A | Internal medicine, allergology | Conservative |
| 10E | Internal medicine, endocrinology | Conservative |
| 10F | Geriatrics | Conservative |
| 10G | Internal medicine, gastroenterology | Conservative |
| 10H | Clinical hematology | Conservative |
| 10I | Infectious diseases | Conservative |
| 10K | Cardiology | Conservative |
| 10M | Nephrology | Conservative |
| 10R | Rheumatology | Conservative |
| 10Y | General internal medicine | Conservative |
| 11 | Anesthesiology and intensive care medicine | Operative |
| 15 | Acute care medicine | Conservative |
| 15E | Acute care medicine, hospital | Conservative |
| 15L | Acute care medicine, hospital, pediatrics | Conservative |
| 15Y | Acute care medicine, primary health care | Conservative |
| 20 | Surgery | Operative |
| 20B | Breast surgery | Operative |
| 20E | Endocrine surgery | Operative |
| 20G | Gastrointestinal surgery | Operative |
| 20H | Pancreatic surgery | Operative |
| 20J | Hand surgery | Operative |
| 20K | Heart transplantation | Operative |
| 20L | Pediatric surgery | Operative |
| 20M | Liver surgery | Operative |
| 20N | Lung transplantation | Operative |
| 20O | Orthopedic surgery | Operative |
| 20P | Plastic surgery | Operative |
| 20R | Heart surgery | Operative |
| 20S | Ambominal transplant surgery | Operative |
| 20T | Thoracic surgery | Operative |
| 20U | Urology | Operative |
| 20V | Vascular surgery | Operative |
| 20Y | General surgery | Operative |
| 20Z | Trauma surgery | Operative |
| 25 | Neurosurgery | Operative |
| 30 | Obstetrics and gynecology | Operative |
| 30A | Obstetrics | Operative |
| 30E | Gynecological endocrinology | Operative |
| 30N | Gynecology | Operative |
| 30Q | Perinatology | Operative |
| 30S | Gynecological oncology | Operative |
| 30U | Gynecological urology | Operative |
| 40 | Pediatrics | Conservative |
| 40A | Pediatric allergology | Conservative |
| 40D | Neonatology | Conservative |
| 40E | Pediatric endocrinology | Conservative |
| 40G | Pediatric gastroenterology | Conservative |
| 40H | Pediatric hematology | Conservative |
| 40I | Pediatric infectious diseases | Conservative |
| 40K | Pediatric cardiology | Conservative |

| | | |
|---|---|---|
| 40M | Pediatric nephrology | Conservative |
| 40L | Pediatric surgery | Operative |
| 40O | Pediatric orthopedics and traumatology | Operative |
| 40P | Pediatric pain medicine | Operative |
| 40S | Pediatric heart and transplant surgery | Operative |
| 50 | Ophtalmology | Operative |
| 50G | Glaucoma surgery | Operative |
| 50H | Oculoplastic surgery | Operative |
| 50K | Cataract surgery | Operative |
| 50M | Retinal diseases | Operative |
| 50N | Neuro-ophtalmology | Operative |
| 50P | Ophtalmologic acute care | Operative |
| 50S | Corneal surgery | Operative |
| 50V | Retinal surgery | Operative |
| 55 | Otorhinolaryngology (ENT) | Operative |
| 55A | ENT, allergology | Operative |
| 55B | Audiology | Operative |
| 55C | ENT, audiology, adults | Operative |
| 55K | ENT, laryngology | Operative |
| 55L | Pediatric otorhinolaryngology | Operative |
| 55O | ENT, otology | Operative |
| 55P | Acute care otorhinolaryngology | Operative |
| 55R | ENT, rhinology | Operative |
| 55T | ENT, malignant diseases | Operative |
| 57 | Phoniatrics | Conservative |
| 57B | Phoniatric audiology | Conservative |
| 58 | Oral and maxillofacial surgery | Operative |
| 58E | Oral infections, primary health care | Operative |
| 58F | Oral infections, hospital | Operative |
| 58T | Oral tumors | Operative |
| 58V | Oral and maxillofacial surgery | Operative |
| 58X | Orthodontics | Operative |
| 58Y | Clinical dental care | Operative |
| 60 | Dermatology | Conservative |
| 60A | Dermatologic allergology | Conservative |
| 60C | Occupational dermatology | Conservative |
| 60I | Dermatology | Conservative |
| 65 | Oncology | Conservative |
| 65W | Rare diseases | Conservative |
| 70 | Psychiatry | Conservative |
| 70A | Acute care psychiatry | Conservative |
| 70C | Mood disorders | Conservative |
| 70F | Geriatric psychiatry | Conservative |
| 70X | Youth psychiatry, psychiatry | Conservative |
| 70Z | Forensic psychiatry | Conservative |
| 74 | Youth psychiatry | Conservative |
| 75 | Child psychiatry | Conservative |
| 75X | Youth psychiatry, child psychiatry | Conservative |
| 77 | Neurology | Conservative |
| 77K | Neurologic rehabilitation | Conservative |
| 77F | Neurologic geriatry | Conservative |
| 78 | Child neurology | Conservative |
| 80 | Respiratory medicine | Conservative |

| | | |
|---|---|---|
| 80A | Respiratory medicine, allergology | Conservative |
| 80X | Lung cancer | Conservative |
| 91K | Clinical pharmacology | Conservative |
| 93 | Sports medicine | Conservative |
| 94 | Genetic medicine and hereditary diseases | Conservative |
| 95 | Occupational and environmental medicine | Conservative |
| 96 | Physical Medicine and Rehabilitation | Conservative |
| 97 | Geriatrics | Conservative |
| 98 | General medicine | Conservative |
| 98T | General medicine, family practice | Conservative |

as an explanatory variable. The model's performance, assessed using 5-fold cross-validation on 46,700 training data points, yielded a root mean squared error (RMSE) of 0.026.

#### S1.4 Patient blood volume

To calculate blood loss relative to blood volume, we mapped patient weight and height data from a separate table. Height and weight at the time of transfusion were selected to be the closest measurements to transfusion start time, with weight measurement being at most 30 days old. If one of weight or height measurements was missing but a BMI measurement existed, the missing variable was calculated using the other two using the BMI formula:

$$\text{BMI} = \frac{\text{weight}}{\text{height}^2},$$

$$\text{height} = \sqrt{\frac{\text{weight}}{\text{BMI}}},$$

$$\text{weight} = \text{height}^2 \times \text{BMI}.$$

We then estimated each patients blood volume using Nadler's equation[2]:

$$V = \begin{cases} 0.3669 \times \text{height}^3 + 0.03219 \times \text{weight} + 0.6041 & |\text{if male} \\ 0.3561 \times \text{height}^3 + 0.03308 \times \text{weight} + 0.1833 & |\text{if female} \end{cases}$$

### S2 Supplementary findings

#### S2.1 Weekly blood use, spanning a year

Figure S2.1 illustrates the relative blood use patterns across different categories over a two-year period (2021-2022). Each data point represents the mean value between corresponding weeks in 2021 and 2022, normalized against the maximum observed weekly mean to facilitate comparison. We found a clear distinction between blood use patterns in elective and urgent operations. Urgent procedures showed relatively consistent blood use throughout the year, with fluctuations. Elective procedures exhibited more pronounced variability, with a marked decrease during the summer months (June-August). This reduction corresponds to the traditional summer holiday period for hospital staff, resulting in fewer scheduled elective surgeries. There was no discernible difference in the overall patterns of blood use between conservative and operative medical specialties. Both exhibited similar fluctuations and relative usage levels throughout the year, suggesting that blood demand is relatively consistent across different medical specialties.

#### S2.2 Time-to-transfusion distributions by ICD-10 category

In Figure S2.2, the x-axis represents the time difference (in days) between the first occurrence of the associated ICD-10 code in the medical records and a transfusion event. The figure demonstrates differences in transfusion timing across different disease categories. For instance, diseases of blood and immune mechanism, pregnancy-related conditions, and digestive system disorders show a pronounced peak near day 1, indicating that transfusions often occur shortly after diagnosis or admission. In contrast, diseases of the circulatory system and musculoskeletal system diseases display a more dispersed distribution, suggesting that transfusions may be required variably depending on the exact diagnosis.

Figure S2.1 | Weekly blood use, spanning a year

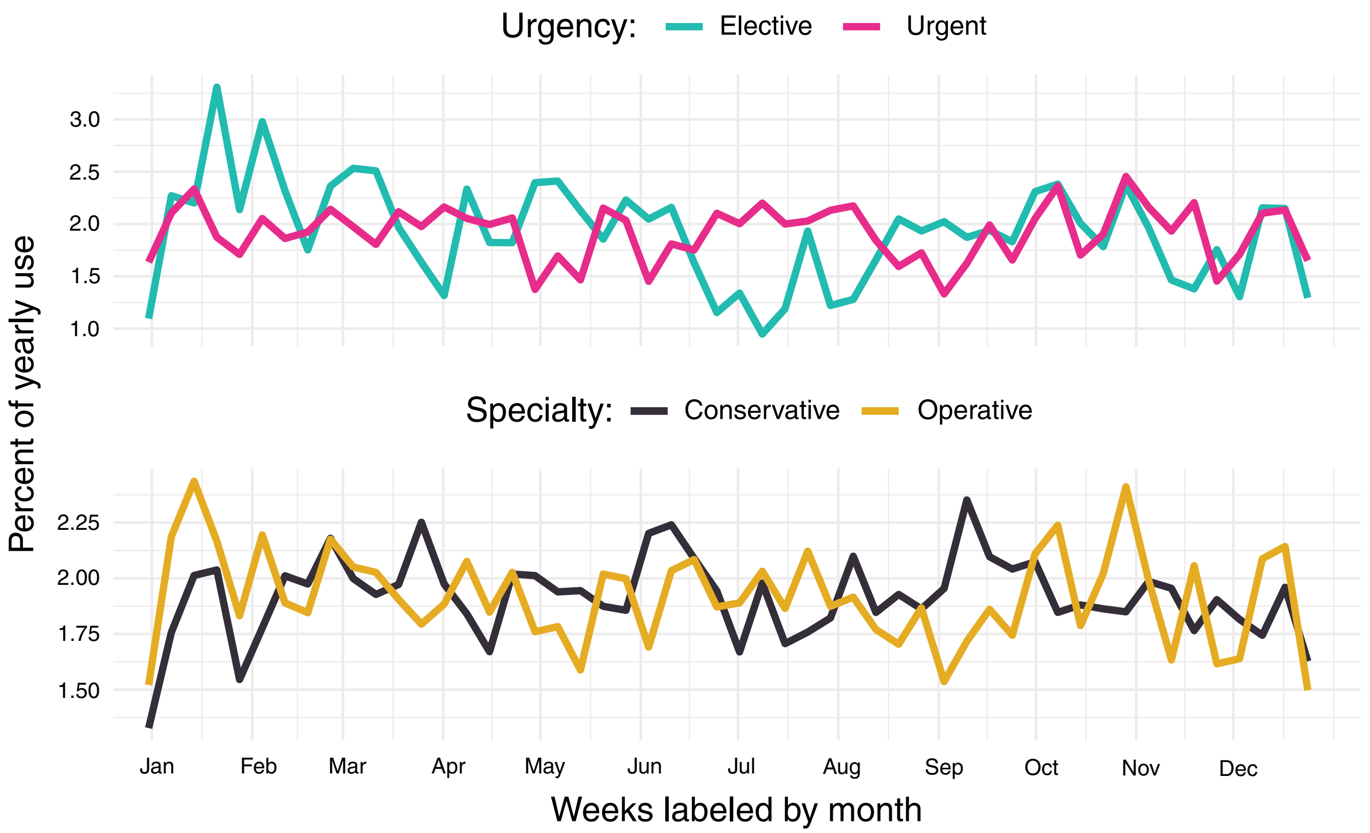


Notably, blood demand for congenital malformations peaks much later, suggesting a complex and long-term disease profile.

### S2.3 Transfused units per transfusion episode

Figure S2.3 presents the distribution of transfused units in distinct transfusion episodes. In most transfusion episodes, only one RBC unit was transfused at a time (including multicomponent transfusions, in which the patient received only one RBC unit concomitantly with other blood units). For platelets, most commonly two units were transfused at a time. For plasma, almost 50% of units were transfused as batches of 8-10 units, indicating use for plasmapheresis or plasma exchange.

### S2.4 Transfused units per treatment episode

For treatment episodes (covering all transfusion episodes associated with one treatment entity for a certain diagnosis), most frequently two RBC units were transfused, indicating that often one treatment episode contains two separate transfusion episodes (Figure S2.4). Similarly for plasma and platelets, two units were transfused most frequently. The distribution of large plasma transfusions is similar to that of transfusion episodes, whereas the number of treatment episodes with large (+5) RBC or platelet transfusions increased compared to the more time-restricted transfusion episodes.

### S2.5 Platelet count and INR pre- and post-transfusion

Distributions of (a) platelet count (x$10^9$/L) and (b) International Normalized Ratio (INR) in transfusion recipients, measured before and after transfusion episodes (Figure S2.5). Only patients with both pre- and post-transfusion measurements taken within a 24-hour interval are included. In addition, we excluded plasma exchange events from the INR distributions. The median transfusion threshold for platelets was relatively high in all patient groups, which can be explained by the case mix including several patient groups with acute bleeding.

Figure S2.2 | Time-to-transfusion by ICD-10 category

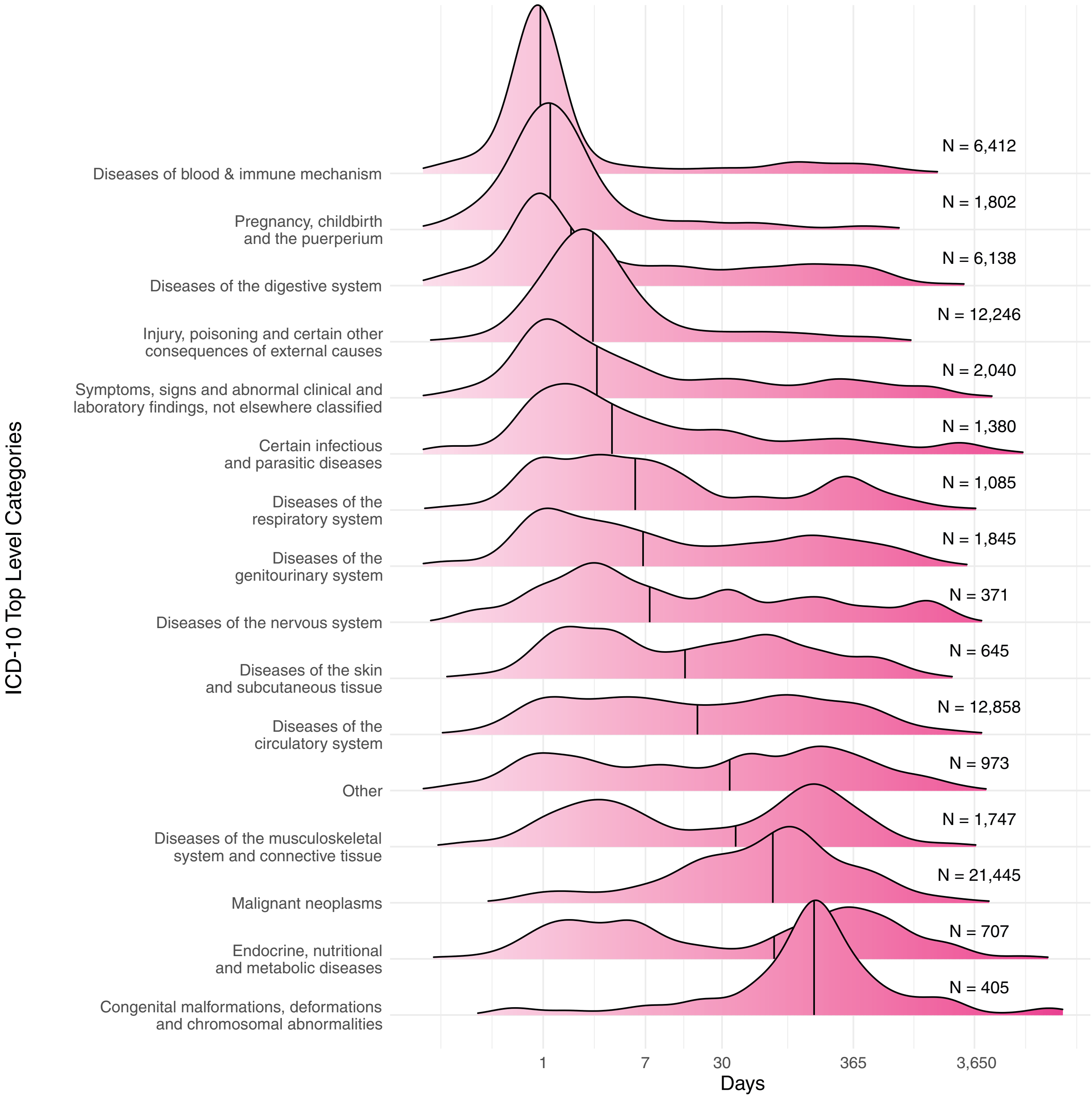


### S2.6 ABO groups of transfused units and patients

Figure S2.6 presents the ABO blood group distribution of transfused units for each patient blood group by unit type.

### S2.7 Time between transfusions

Figure S2.7 presents the distributions of time between the end of a patient's transfusion and the start of their next transfusion, by unit type. While the long tails of the distributions indicate that many patients receive their sequential transfusions with significant delay, most units are transfused within the set 3-hour limit for a single transfusion episode.

### S2.8 Patients receiving RBC units at a higher pre-transfusion Hb limit

Both in men and postmenopausal women, pre-transfusion Hb distribution indicated two distinct transfusion thresholds: 70 to 79 g/L (peak in the distribution) and 80-89 g/L ("shoulder" in the distribution) (Figure 8). We compared these with respect to patient age, main diagnosis, and main procedure. Figure S2.8 presents the proportion of transfusions given with a higher (80-89 g/L) pre-transfusion Hb versus

### Figures S2.3 & S2.4 | Transfused units per transfusion episode and treatment episode

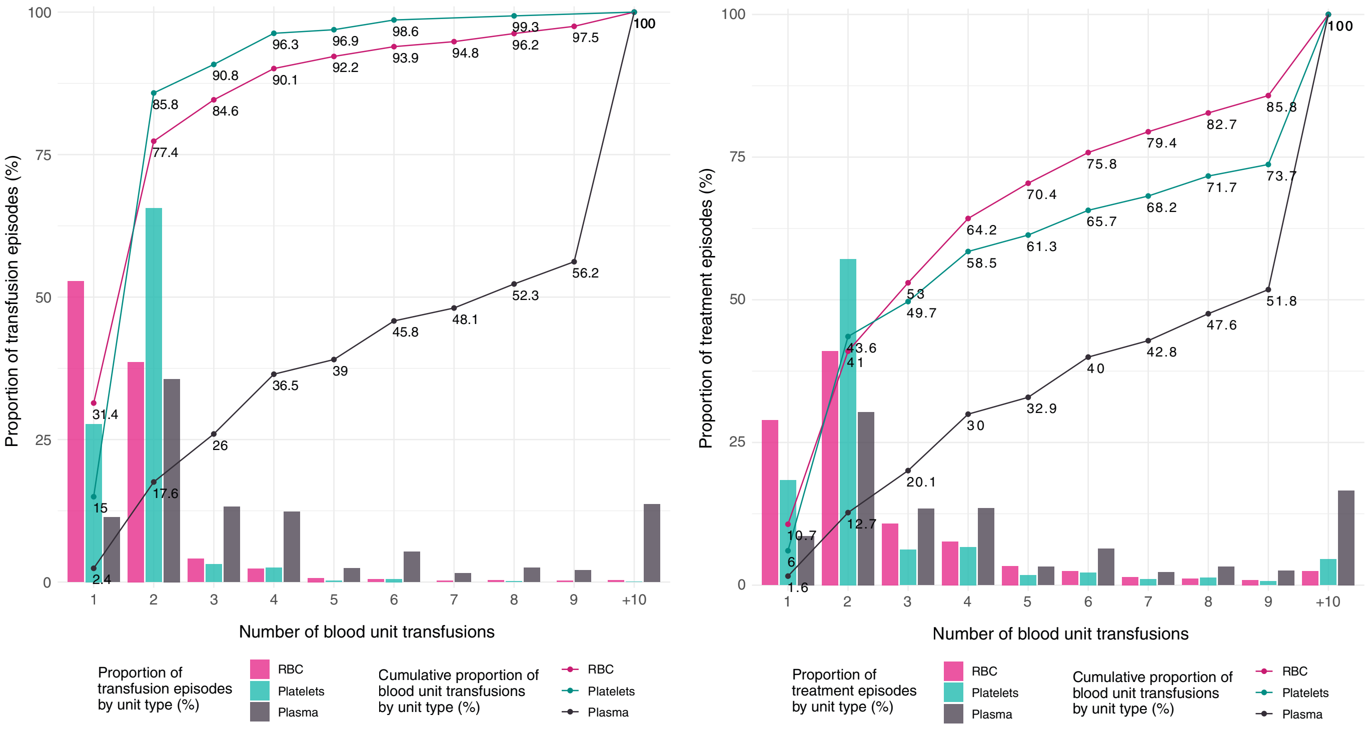


### Figure S2.5 | Patient platelet count and INR before and after transfusion episode

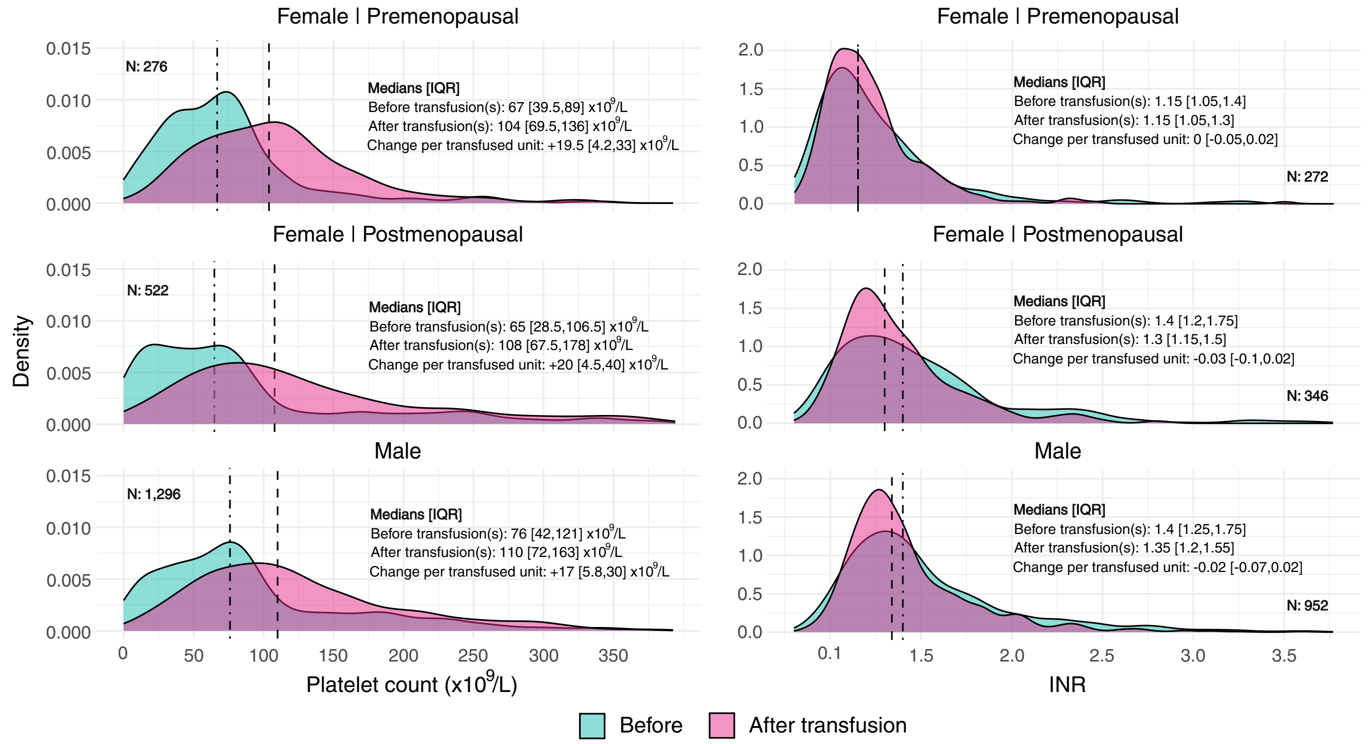


lower (70-79 g/L) by patient sex and age groups. We excluded premenopausal women from this analysis, as their pre-transfusion Hb distribution did not indicate distinct transfusion thresholds. Table T2.8 lists the ICD-10 categoriers and functional-anatomical regions from the NCSP that are most disproportionately associated with a higher pre-transfusion Hb (80-89 g/L). Skew is presented as the factor of overrepresentation (e.g. a skew of 2 represents the ratio of 1:2). Skews were computed only for common diagnoses (>500 per sex group) and procedures (>200 per sex group), as rare diagnoses and procedures might skew much more to higher pre-transfusion Hb levels, but would not contribute enough to the formation of the "shoulder" in the distribution.

### Figure S2.6 | ABO group of transfused units and patients

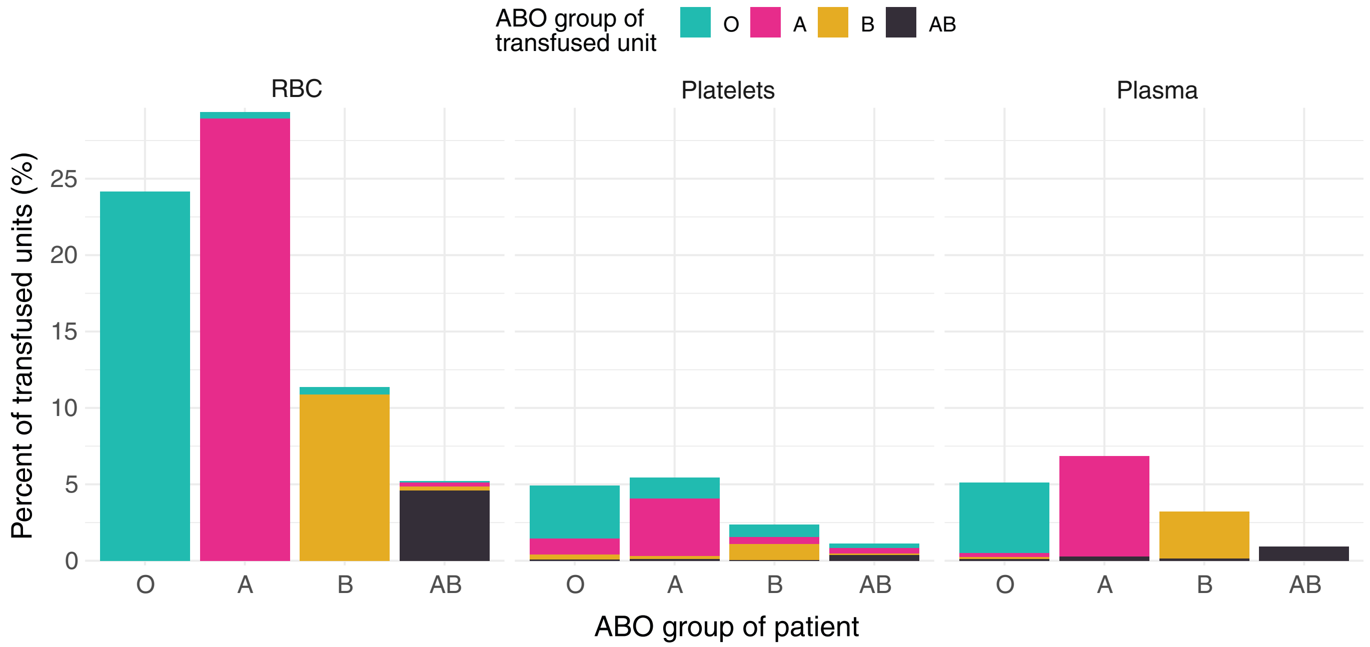


### Figure S2.7 | Time between units transfused by unit type

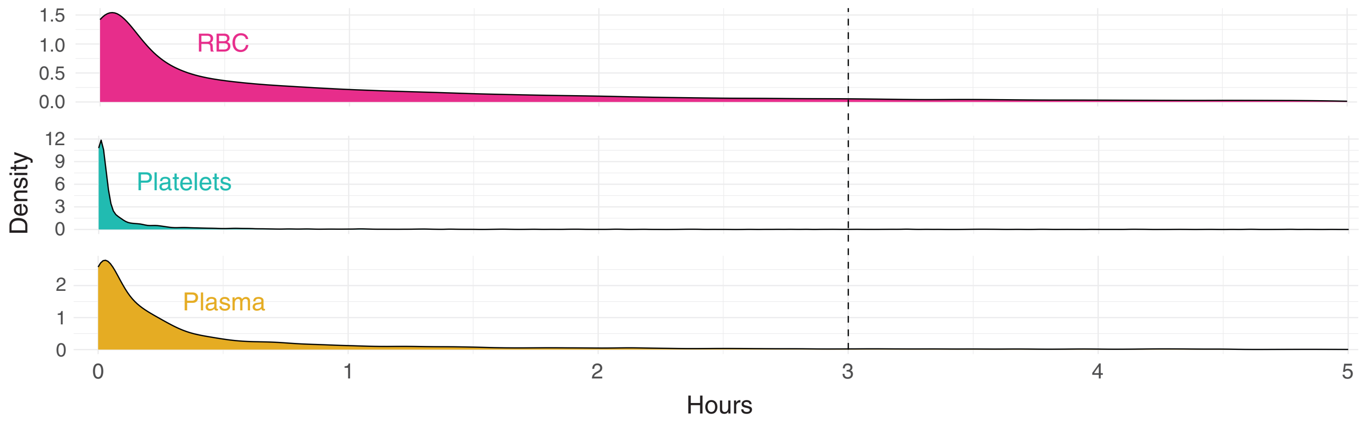


### Figure S2.8 | The proportion of transfused units at pre-transfusion Hb levels of 80-89 g/L vs. 70-79 g/L by age groups

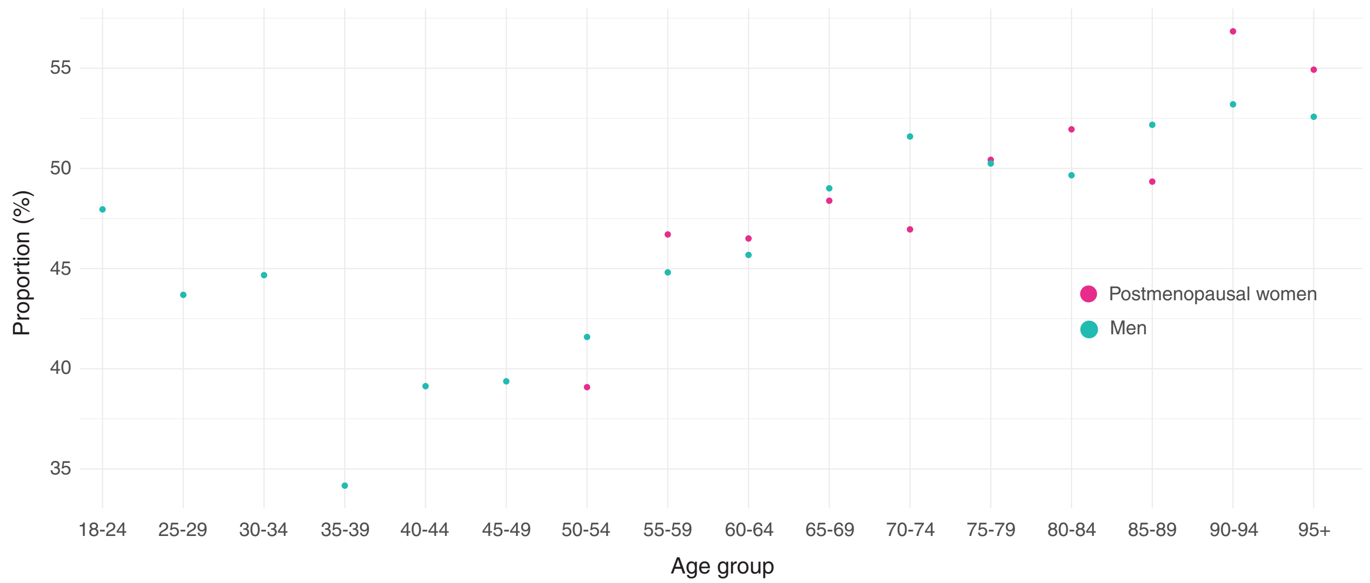

Table T2.8 | ICD-10 categories and NCSP functional-anatomical regions most over-represented in RBC transfusions at pre-transfusion Hb levels 80-89 g/L vs. 70-79 g/L

| ICD-10 Category | Skew | N |
|---|---|---|
| **Female, postmenopausal** | | |
| I70-79 \| Diseases of arteries, arterioles and capillaries | 2.41 | 628 |
| T80-88 \| Complications of surgical and medical care, not elsewhere classified* | 1.41 | 778 |
| S70-79 \| Injuries to the hip and thigh | 1.33 | 1,918 |
| **Male** | | |
| I70-79 \| Diseases of arteries, arterioles and capillaries | 1.78 | 1,289 |
| S70-79 \| Injuries to the hip and thigh | 1.65 | 831 |
| T80-88 \| Complications of surgical and medical care, not elsewhere classified* | 1.59 | 1,144 |

| NCSP Category | Skew | N |
|---|---|---|
| **Female, postmenopausal** | | |
| PE \| Femoral artery with branches and connection to popliteal artery | 2.17 | 219 |
| NG \| Knee joint and lower leg | 1.91 | 405 |
| NF \| Hip joint and thigh | 1.36 | 2,238 |
| **Male** | | |
| PE \| Femoral artery with branches and connection to popliteal artery | 2.51 | 260 |
| NF \| Hip joint and thigh | 1.95 | 1,139 |
| KC \| Bladder | 1.38 | 202 |

*Diagnosis code most commonly associated with the following reoperations: revision arthroplasty, postoperative bleeding, surgical site infection, bowel anastomotic leak, vacuum-assisted or delayed wound closure.
